# Social distancing and movement constraint as the most likely factors for COVID-19 outbreak control in Brazil

**DOI:** 10.1101/2020.05.02.20088013

**Authors:** Matheus Tenório Baumgartner, Fernando Miranda Lansac-Tôha, Marco Túlio Pacheco Coelho, Ricardo Dobrovolski, José Alexandre Felizola Diniz-Filho

## Abstract

As thousands of new cases of COVID-19 have been confirmed, there is an increasing demand to understand the factors underlying the spread of this disease. Using country-level data, we modeled the early growth in the number of cases for over 480 cities in all Brazilian states. As the main findings, we found that the percentage of people respecting social distancing protocols was the main explanatory factor for the observed growth rate of COVID-19. Those cities that presented the highest spread of the new coronavirus were also those that had lower averages of social distancing. We also underline that total population of cities and connectivity, represented by the city-level importance to the air transportation of people across the country, plays important roles in the dissemination of SARS-CoV-2. Climate and socioeconomic predictors had little contribution to the big-picture scenario. Our results show that different States had high variability in their growth rates, mostly due to quite different public health strategies to retain the outbreak of COVID-19. In spite of all limitations of such a large-scale approach, our results underline that climatic conditions are likely weak limiting factors for the spread of the new coronavirus, and the circulation of people in the city- and country-level are the most responsible factors for the early outbreak of COVID-19 in Brazil. Moreover, we reinforce that social distancing protocols are fundamental to avoid critical scenarios and the collapse of healthcare systems. We also predict that economic-induced decisions for relaxing social distancing might have catastrophic consequences, especially in large cities.

## 1. Introduction

In late December, 2019, the novel Coronavirus Disease 2019 (COVID-19) emerged in Wuhan, the capital city of Hubei Province in great China (Zhu et al., 2020). At the beginning of the infection outbreak, the disease caused by the SARS-CoV-2 virus has been suggested to be of bat origin (Cheng et al., 2007; Guo et al., 2020; Zhou et al., 2020), and might have been transmitted to humans through intermediate mammals (Andersen et al., 2020; Li et al., 2020; Zhang et al., 2020). The initial diffusion of the virus suddenly became exponential, increasing the number of infected cases and deaths in Wuhan (Kraemer et al., 2020). In the next few weeks, it has become clear that the high virulence of the new coronavirus posed a considerable health threat on a global scale, quickly spreading to Asia, Europe, North America, and, more recently, South America and Africa.

On March 11, 2020, the World Health Organization (WHO) declared the coronavirus as a global pandemic. The declaration reflected the concern by WHO that countries were unable to control the dissemination of the virus (Li et al., 2020). The novel coronavirus is still spreading rapidly worldwide, with millions of infections and hundreds of thousands deaths addressed to COVID-19, most of them concentrated in the United States and Europe.

Common symptoms of COVID-19 infections include fever, dry cough, and dyspnea but the most serious clinical case is lung failure associated with severe acute respiratory syndrome (SARS) (Yang et al., 2020). From all infected patients, about 30% require mechanical ventilation and 2-5% die, with higher death rates in elderly people and those with comorbidities (Rothan and Byrareddy, 2020). Fortunately, most people have very mild symptoms or are even asymptomatic (Yang et al., 2020).

Conventionally, laboratory testing by real-time polymerase chain reaction (RT-PCR) and quick antigen tests have been conducted prioritizing symptomatic or high-risk groups (Mizumoto et al., 2020). However, a recent study showed that substantial undocumented infection facilitates the rapid dissemination of the novel coronavirus (R. Li et al., 2020), which forced the WHO to recommend that policymakers should mobilize mass testing in an attempt to retain initial local outbreaks effectively (Balilla, 2020).

With clinical symptoms that are indicative of many ordinary conditions, the new coronavirus is the largest concern for human populations because it may cause severe clinical conditions that can readily overcome the carrying capacity of healthcare facilities. Although vaccines and immunotherapy protocols have been conducted in a rate never seen before, there is no effective treatment for COVID-19 yet (Lurie et al., 2020). The available interventions include rapid diagnosis and isolation of confirmed cases, and restrictions on mobility (Kraemer et al., 2020). Especially for in-development countries, social isolation has been listed as the most effective strategy to deal with COVID-19 and mitigate the risk for public health and economy (Coelho et al., 2020). However, understanding how other environmental and social factors are associated with human-to-human transmissions of SARS-CoV-2 are fundamental for the decision-making process at a country-level.

Building on evidence about the role of environmental factors such as temperature and humidity on the survival of viruses, some studies forecasted the near future of the current outbreak (Sajadi et al., 2020; Wang et al., 2020). For instance, Araújo and Naimi (2020) built a global ensemble to model the monthly spread of COVID-19 under the prediction of temperature and humidity, although with some later criticism (Chipperfield, 2020). While these variables are known to interfere with the spread and survival of other coronaviruses (e.g., SARS-CoV and MERS-CoV; Gaunt *et al*., 2010; Chan *et al*., 2011; Cauchemez *et al*., 2014), considering only the environment and ignoring social and behavioral factors might be inadequate to determine effective restraint strategies in the near future (Pybus et al., 2015).

Considering socioeconomic aspects, countries and regions across the world have a very skewed distribution of income and wealth (Dabla-Norris et al., 2015; Davies et al., 2017). Even within countries, there are clear spatial distribution patterns of economic development, as it is explicit within Brazil (Skidmore, 2004). In terms of healthcare, socioeconomic indicators can be considered as a proxy for the ability of each city to identify and treat people with COVID-19 effectively (Coelho et al., 2020). Additionally, the dispersal of viruses among hosts follows a geographic pattern (Holmes, 2004). Thus, the physical distance among cities (and people) is certainly crucial for how the actual pandemic state will evolve (Chipperfield, 2020). In an attempt to investigate these factors, Coelho et al. (2020) found evidence that the air transportation of people across the word overcame environmental and socioeconomic factors, posing a strong argument towards social distancing and traveling restraint.

Thus, here we extended the investigation on which factors could be related to the spread of the new coronavirus in Brazil, considering all cities on a nation-wide scale. We studied the exponential growth of time series data for over 460 cities with reported cases of infections by the new coronavirus, considering the effect of the environment, socioeconomic indicators, movement of people across the country, and social distancing. We demonstrate that the growth of COVID-19 in different cities is mostly determined by population size, transportation among cities, and the percentage of people respecting social distancing protocols. This evidence points towards social distancing and mobility restriction as the main actions necessary to reduce the spreading of the pandemic and avoid its worst consequences.

## 2. Material and Methods

### 2.1 Dataset and exponential growth model

We obtained data on the daily number of people manifesting the COVID-19 in Brazil, available at the digital panel from the Ministry of Health (http://www.covid.saude.gov.br). This dataset comprises real-time information on disease cases that were confirmed through laboratory analysis and quick tests for every city. Our most recent data retrieval was conducted on April 20, 2020, which comprised information of 40,581 cases and time series data for 1,456 cities with at least one confirmed case, since the first recorded case in Brazil on February, 25, 2020. We chose April 20 as the last day because several cities have gradually relaxed their quarantine protocols and allowed many working activities to restore their functioning since then. In our analyses, we used only the exponential part of each time series by excluding previous days before the first confirmed cases in each city, and no time series had reached stabilization or decrease in the total number of confirmed records yet.

We fitted exponential growth models to the time series of each city and calculated the intrinsic growth rate (*r*), as well as the slope coefficient (*b*) of the log-growth model. Because our focus was on the overall growth of the number of confirmed cases, these two parameters were candidates to be used as response variables in our approach. However, some convergence failures for exponential models applied on short time series and the high goodness-of-fit of log-transformed models (average *R*^2^ = 0.84) led us to use the log-growth slope in our subsequent analysis.

### 2.2 Predictor variables

In order to explore potential correlates of the early increase in the number of cases, we used climatic and socioeconomic data. Climatic variables included average temperature (°C) and precipitation (mm), retrieved from the most recent year available at the WorldClim online database (http://www.worldclim.org; Fick & Hijmans, 2017). This database comprises monthly information on these climatic variables. We downloaded temperature and precipitation values between February and April, which coincided with the time series of COVID-19 cases in Brazil and with the late summer season of the Southern Hemisphere, matching the world outbreak of the virus. From these data, we extracted values for each city according to their geographic coordinates, using the QGIS 3.12.1 software (QGIS Development Team, 2020).

For each city, we also extracted information on total population, Human Development Index (HDI), and average income. These demographic data were obtained from the digital platform maintained by the Institute of Applied Economic Research, under the ‘*Atlas do Desenvolvimento Humano no Brasil*’ project (http://www.atlasbrasil.org.br/). This initiative encompasses data about more than 5,500 municipalities in all 27 Brazilian federal units, with several socioeconomic descriptors derived from the 2010 national census.

As an additional correlate to the increase of the infestation by the new virus, we also considered information on transportation of people across the country. First, we obtained data on air transportation available at the OpenFlights database (http://openflights.org/data.html), which included information on 122 airports within Brazil, and whether there is a direct flight connecting each pair of them (1,193 flights). To match the information on the spread of the virus across cities, we filtered these airport records to consider only those cities that had at least one confirmed infection, which yielded us a database containing 100 cities with 1,164 registered flights (Fig 1). We then used these cities and flights to construct an oriented graph with cities assigned as notes and flight routes as edges (West, 2001). From this network, we extracted a variable that weighted the importance of each city to the country-level network: the Eigenvector Centrality (Bonacich, 1987). This metric quantifies both how surrounding cities are connected to a focal city and its connection to the whole network, directly and indirectly. Second, many cities with confirmed cases are not large enough to carry airports with commercial flights. In these cases, we used the geographic coordinates to calculate the proximity to the nearest airport (1 - standardized Euclidean distances), which was then multiplied by the centrality of their respective nearest airport. These two procedures yielded connectivity, a variable that represented the direct and indirect movement of people among large cities, based on their centrality, as well as their potential exchange of people with smaller municipalities nearby, described by the proximity-weighted centrality.

**Fig. 1:**
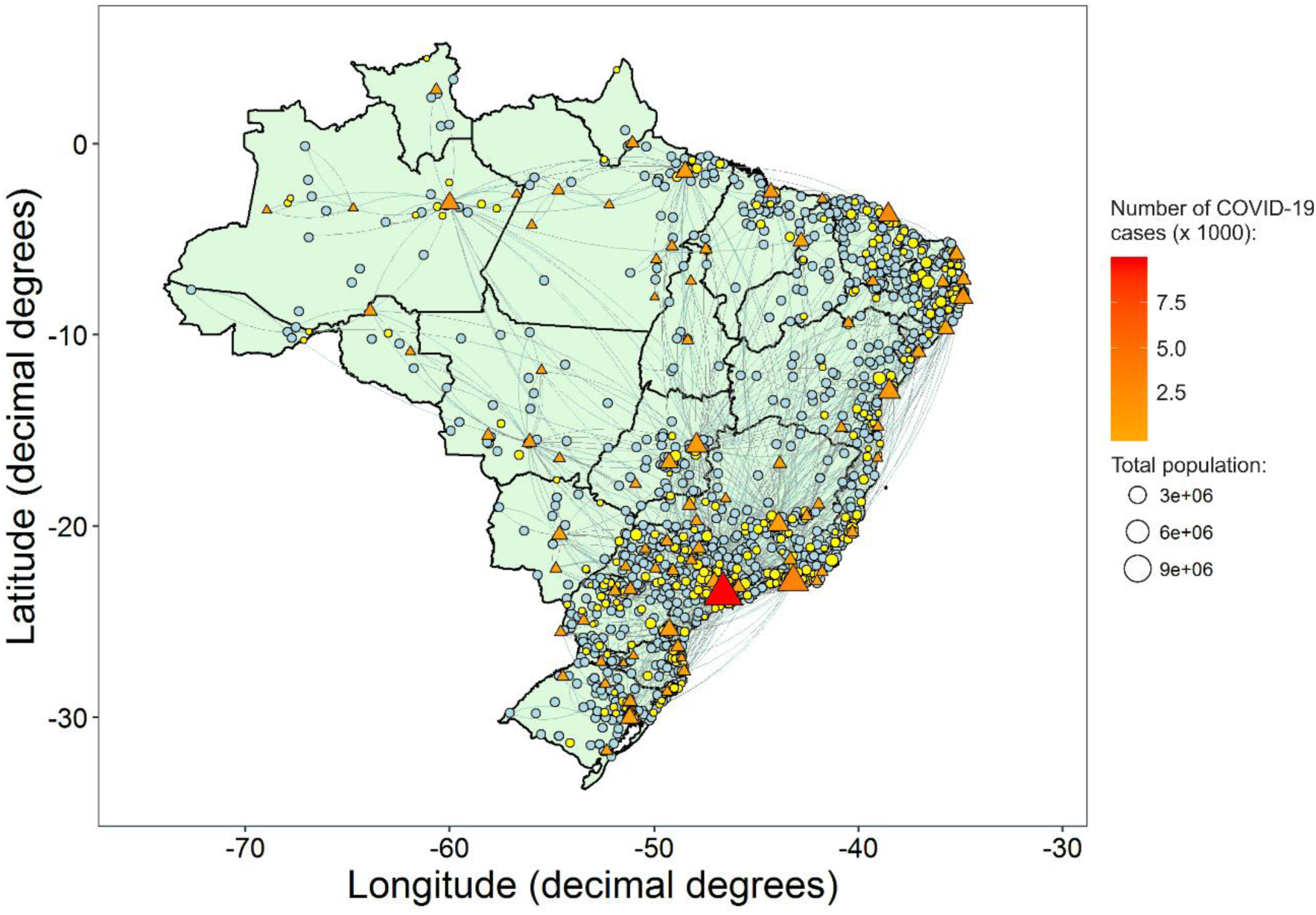
Geographic distribution of 1,456 cities with at least one confirmed case of COVID-19 in Brazil by April 20, 2020. Blue circles represent cities with less than five confirmed cases, which were not included in our analyses. Yellow, orange, and red symbols depict cities that had at least five confirmed cases, with (82 cities; triangles) and without (383 cities; circles) airports. Flying routes are depicted by gray traces and State borders are in black. See Fig. S1 for more details on the names and locations of each Brazilian state.

In order to assess the effectiveness of social distancing on the early spread of the new coronavirus in Brazilian cities, we used State-level data on mobility of citizens (https://mapabrasileirodacovid.inloco.com.br/), which comes from a monitoring program that was exclusively implemented to be useful against the new coronavirus. This database comprises the percentage of people performing social distancing for each state on a daily basis. The index is based on information provided by telephone companies about the location of electronic devices (e.g., cellphones and tablets), which are tracked through their physical displacement under Wi-Fi, Bluetooth, and GPS connections. In practice, whenever an electronic device leaves a radius of ~200 m from what it is considered as ‘home’, the system records the event as a movement. Therefore, this protocol ensures the privacy and preserves the identity of citizens by focusing on the binary record of movement, rather than on the destination or specific routes of each citizen. This information yielded us a database with the time series of social distancing for each state. To obtain city-level data, we averaged the percentages between the first day of COVID-19 record in Brazil and the day where the thresholds of the minimum number of cases was achieved at each city (see below). See Supplementary Material Fig. S1 for more details on the names and locations of each Brazilian state. Time series of percentages of people respecting social distancing protocols at each Brazilian state are given in Fig. S2.

### 2.3 Data analysis

We investigated whether the city-level growth rate of COVID-19 was related to the climatic, socioeconomic, connectivity, and social distancing predictors using multiple linear regression models. Models were fitted using sequential subsets of the 465 cities considering increasing thresholds of the minimum number of confirmed cases: 5, 10, 20, 40, 80, and 160. This sequential sub-modelling approach intended to assess whether and how the COVID-19 outbreak across Brazil was related to each one of our predictors along the early spread of the disease, considering cities at different stages of the dissemination wave. All predictors were checked and only total population required log transformation to approximate a normal distribution. Before statistical analyses, we checked the multicollinearity of predictors, by computing the variance inflation factors (VIF), and removed those that the variance of a regression coefficient was inflated in the presence of other explanatory variables (i.e. VIF > 5; (Borcard et al., 2018)). In this case, HDI and average income showed collinearity, so we kept the later in our analyses because of its larger variation. In addition, we used Moran’s I correlograms (Legendre and Legendre, 2012) to check if the control for spatial autocorrelation bias was required, which could somehow inflate the significance of each predictor. A summary of all the predictors used to fit the regression models is provided in Fig 2.

Finally, we fitted independent linear models to each State to investigate potential regional trends in the COVID-19 growth, using the most important predictors identified in the previous steps, separately. For this procedure, we used only those states that had at least five cities with at least five confirmed cases. For inference, we then plotted the log-transformed total number of confirmed cases against the standardized slopes of the models, which portrays the state-dependent context of growth in COVID-19 cases. All analyses were performed in the R Environment (R Core Team, 2019) using packages *ape* (Paradis and Schliep, 2019), *car* (Fox and Weisberg., 2019), and *igraph* (Csardi and Nepusz, 2006).

**Fig. 2:**
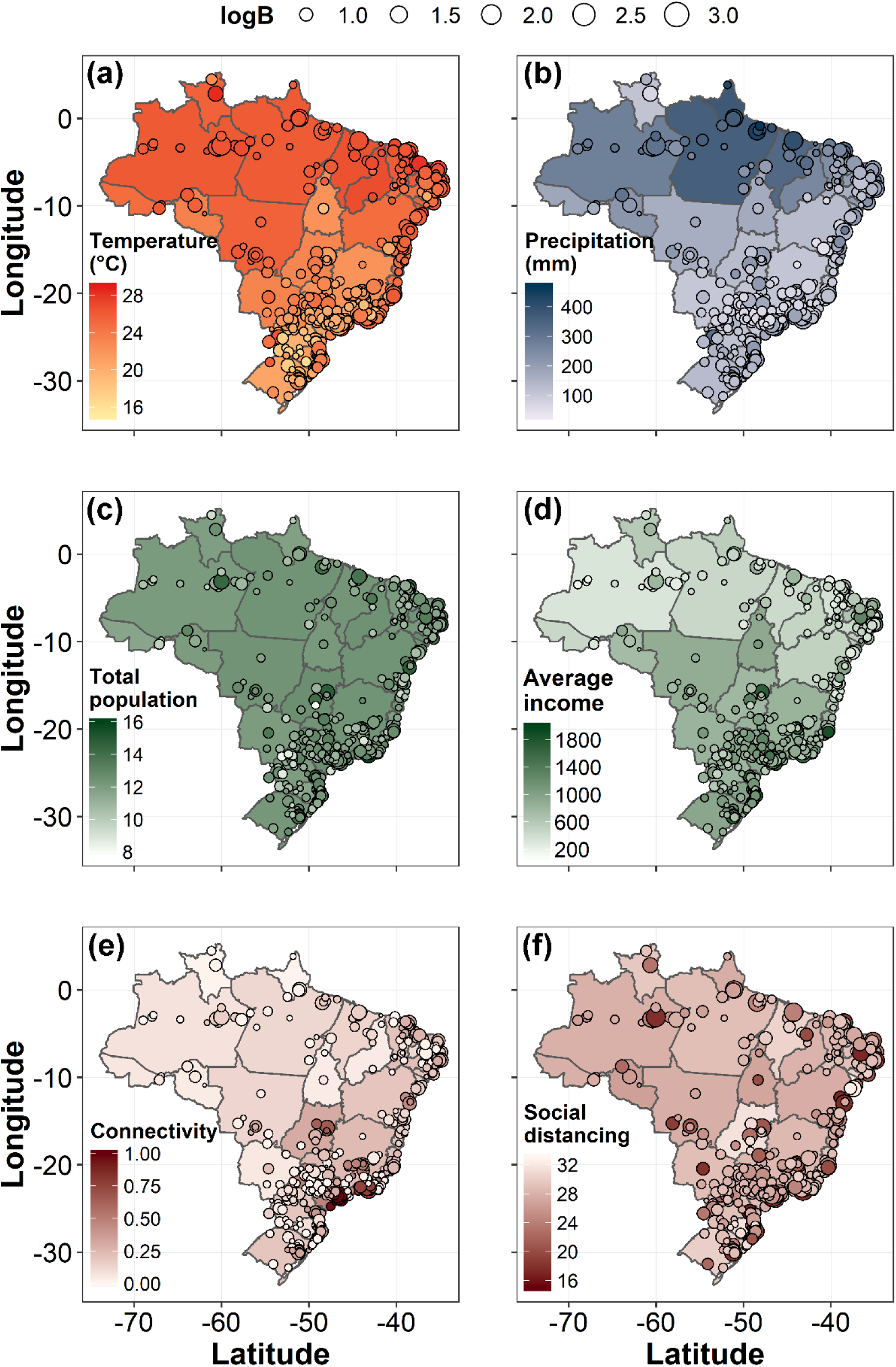
Brazilian maps showing the geographic patterns of all predictors considered in the regression models: temperature (a), precipitation (b), log-total population (c), average income (d), connectivity (e), and social distancing (f). States borders are depicted by black traces and colors of each State represents the mean value considering all cities. Circles depict cities that had at least five confirmed cases. The colors within circles represent the values of each predictor, whereas the circle size depicts the b log growth rate (logB) for each city.

## 3. Results

The overall performance of models fitted to the exponential growth of COVID-19 in each city was particularly good. The average variance explained by our predictors (*R*^2^) were 0.51 (>= 5 cases; 465 cities), 0.60 (>= 10 cases; 252 cities), 0.61 (>= 20 cases; 158 cities), 0.64 (>= 40 cases; 104 cities), 0.56 (>= 80 cases; 62 cities), and 0.67 (>= 160 cases; 33 cities). Considering the effect of each predictor on the exponential growth across all sub-models, social distancing had significant relationships in all cases, with an average standardized coefficient of -0.15 (Table 1, Fig 3). This variable was also the only with negative significant slope coefficients. Most importantly, social distancing had an intensifying trend in its negative coefficient across sub-models, showing an enhanced effect as the number of confirmed cases evolved. In addition, total population and connectivity of each city had significant relationships in five out of six sub-models, with average standardized coefficients of 0.14 and 0.07, respectively. Another predictor that played a secondary role was precipitation, which was significant in the first four sub-models, although with a considerably weak average standardized coefficients (0.05). Average income was only significant for the first sub-model, whereas temperature was not significant in any sub-model. None of the six sub-models presented bias related to spatial autocorrelation structures according to Moran’s I correlograms.

**Table 1.**
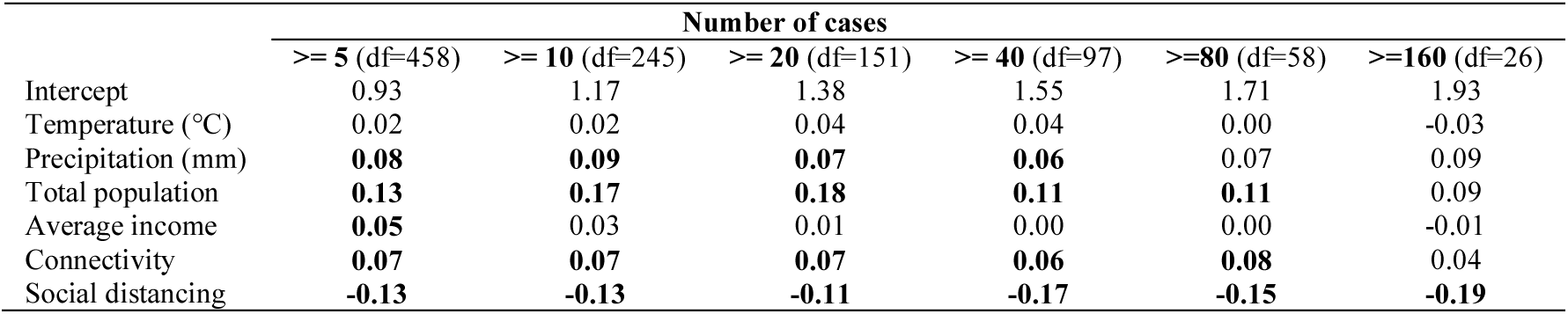
Parameter estimates (standardized *β*‘s) for all variables used as predictors of the slope coefficient (*b*) of the log-growth models, fitted for the number of confirmed cases of infection by Coronavirus Disease (COVID-19). Significant estimates (at *α* = 0.05) are in bold; see Table S1for full results.

**Fig. 3:**
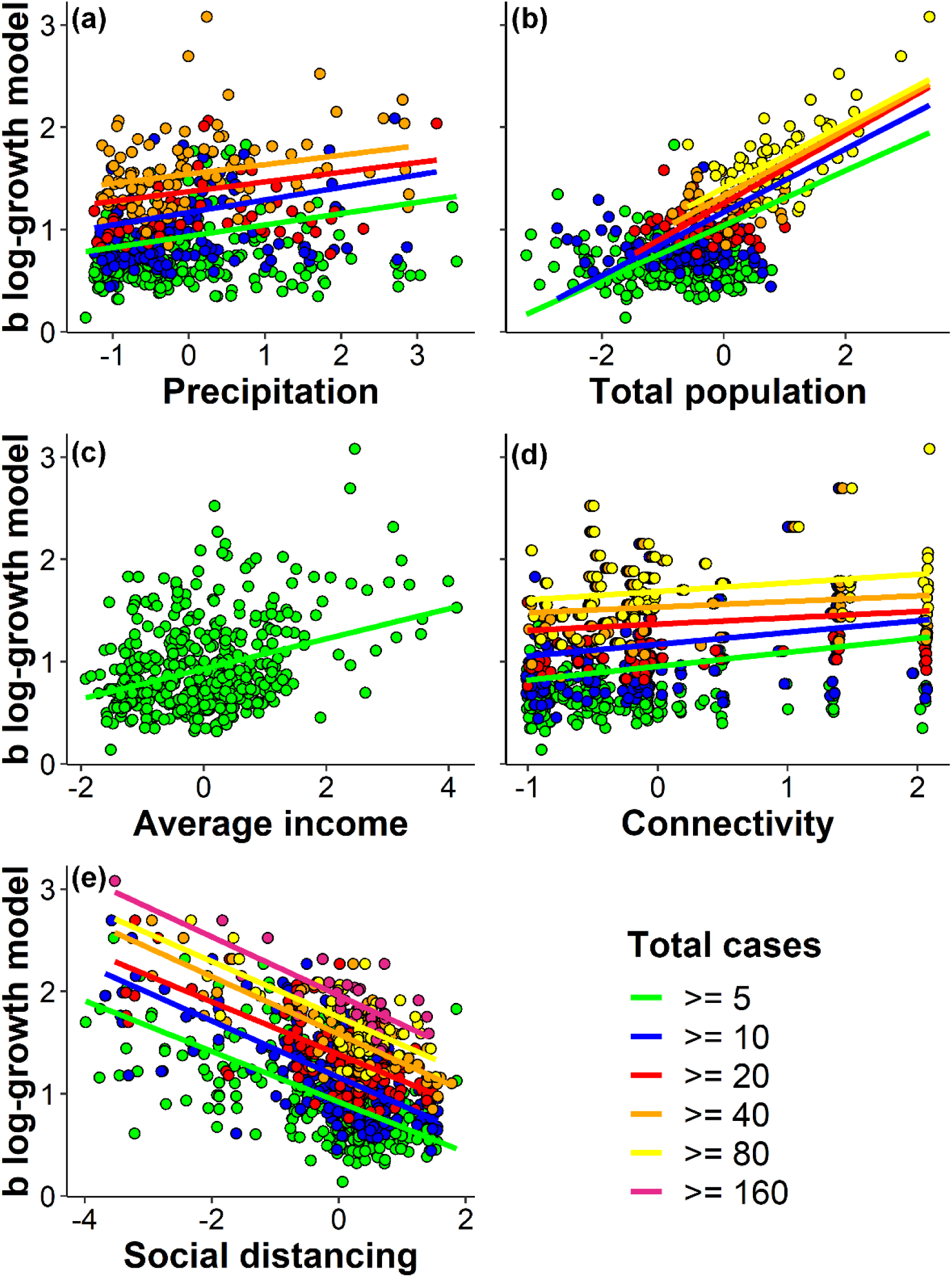
Relationships between the slopes of log-growth models fitted to time series of confirmed COVID-19 cases for every city. Only significant regression models are shown.

Comparatively, the variable with the clearest effect on the exponential growth during the COVID-19 early outbreak was social distancing (Table 1; Fig 3e). In practice, this result suggest that the more frequent people circulated within cities (lower social distancing), the faster the early spread of the new coronavirus (a result that was consistent across all sub-models). The results also reveal that the log-growth of the disease caused by the SARS-CoV-2 virus increased with the total population of cities (Fig 3). Thus, larger population sizes led to faster spread of the new coronavirus. The third most important variable, connectivity (Fig 3d), had a similar trend but with considerably lower standardized coefficients (at least two times lower than total population, Table 1). In contrast, we observed that precipitation (Fig 3a) showed trends that were more important during the initial days in each city, with positive but very low coefficients (Table 1).

When we built one independent model for each State, considering total population, connectivity, and social distancing (i.e., the three most important predictors), there was a high variation in the results depending on the focal State (Fig 4). Notably, the rate of increase in the number of COVID-19 cases was mostly determined by respecting social distancing protocols (i.e., higher R^2^; Fig 4c), followed by population size (Fig 4a). Although not with the same intensity, connectivity also had an important role on the observed growth patterns (Figure 4b).

**Fig. 4:**
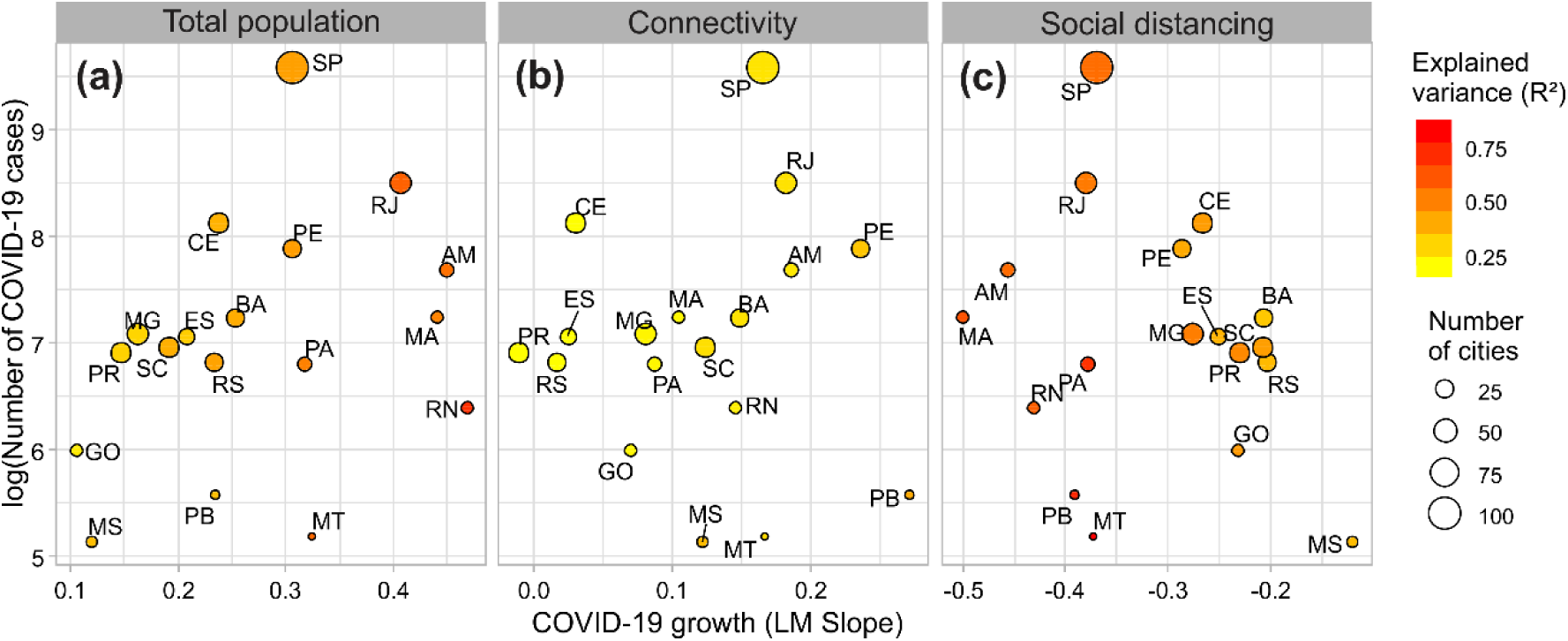
Relationship between the log-transformed total number of confirmed cases against the standardized slopes of the models. (a) total population, (b) connectivity, and (c) social distancing for 18 Brazilian states. See Fig. S1 for more details on the names and locations of each Brazilian state.

The overall trend was that those states where people were performing social distancing less effectively (leftward circles in Fig 4c) coincided with those states under the most positive effect of total population and connectivity (rightward circles in Figures 4a,b, respectively) on the spread of COVID-19. These examples include Amazonas (AM), Maranhão (MA), Rio Grande do Norte (RN), and Rio de Janeiro (RJ). Moreover, total population and social distancing shared similar slices of explained variance (i.e., similar color in Figures 4a,c), such as in Amazonas (AM) and Maranhão (MA). Where the number of cases were the highest in Brazil, such as São Paulo (SP), Rio de Janeiro (RJ), Ceará (CE), and Pernambuco (PE), both population size and social distancing shared nearly similar contributions for their observed condition of the spread of COVID-19. Particularly, the situation of Pernambuco (PE) can be considered as the fairest tradeoff between population size and distance of citizens (i.e., similar circle colors). By contrast, those states where the social distancing and the effect of total population were combined to produce the mildest scenarios of COVID-19 spread were Espírito Santo (ES), Goiás (GO), Mato Grosso do Sul (MS), Paraná (PR), and Santa Catarina (SC), compared to other federal units.

## 4. Discussion

In addition to the pandemic state and the thousands of deaths caused by COVID-19 worldwide, some government boards and leaders manifest that their foremost concern regards the economic retraction, which is expected to last longer than the harmful effects of the COVID-19 outbreak themselves. In fact, they are not completely controversial from their perspective. While in-development countries’ systems depend on economic income (e.g., sales of commodities), further projections are certainly contingent upon the creation and offer of jobs to remain sustainable. However, the transmission of the virus and the complications arising from severe illness conditions may never rest on the bottom of the country-level priorities of one’s management actions. By contrast, decision-makers should provide raw information to fuel research and development of effective containment and treatment of diseases. Hereafter we discuss our results in light of the situations that other countries and cities around the world have experienced.

Our results show that the early spread of the new coronavirus in Brazil was mitigated by social distancing in some regions, but was also positively related to the size of the population of cities and how people moved across them. These outcomes underline that the direct and indirect contact among individuals was the most responsible for the rapid spread of the disease caused by the SARS-CoV-2 virus in Brazil. Contrary to initial perspectives, the ability of COVID-19 to spread, estimated by the basic reproduction number (R0) statistic, seems to be higher than the WHO estimated (Liu et al., 2020). Moreover, substantial transmission before symptom onset facilitates the rapid dissemination of the novel coronavirus (Hellewell et al., 2020). As diseases transmitted by respiratory droplets require a certain proximity of people, social distancing certainly reduce transmission rates (Anderson et al., 2020). For this reason, keeping people apart from each other, whenever feasible, should be the primary goal of public health programs to prevent human-to-human transmission. We found evidences that the tools adopted by states and municipalities, including the closure of schools and commercial buildings, lock down of restaurants and malls, and prohibitions of mass gatherings had a negative impact on the expansion of COVID-19 in Brazil. Most importantly, this result somehow evidences that the combination of ignoring social distance especially in large and well connected cities will have catastrophic consequences.

In Great China, the ongoing COVID-19 outbreak expanded fast throughout the country and the majority of early cases reported outside of its origin had admitted recent travels to Wuhan, the core of the disease spread (Chinazzi et al., 2020). Because of the association between both international and domestic air travel and the dissemination of COVID-19 (Bogoch et al., 2020; Zhao et al., 2020), one of the initial plans for the contagious control was to prevent people from flying around when the outbreak emerged (Kraemer et al., 2020). Actions such as reducing human mobility by restricting travels and declaring quarantine were fundamental to reduce the dissemination of SARS-CoV-2 within and outside Wuhan, highlighting the importance of mobility restrictions in cities where there is a clear potential for spread of the new coronavirus (Fang et al., 2020). Fortunately, in China, the government-level positions were declared when contagious boundaries were relatively discrete (Kraemer et al., 2020).

Comparatively, this was not the case in Brazil, where the number of confirmed cases of COVID-19 are still growing exponentially (Crokidakis, 2020). The first case in Brazil was registered on February 25, 2020, in São Paulo City (Rodriguez-Morales et al., 2020). It was a man with recent travel to the Italy, precisely for the Lombardy region. Contrary to China, Italy did not conduct a fast protocol of social isolation and already had a high number of cases and deaths. Similarly to the observed in the United States, until the establishment of local transmission in Brazil, all reported cases had returned from recent travels abroad. Among them, it is noteworthy that 23 members of the delegation that accompanied the Brazilian President on a visit to the United States in March had since tested positive for COVID-19. Indeed, the proportion of estimated imported cases by airport of destination is highly correlated with the proportion of detected imported cases (Candido et al., 2020). For this reason, the Brazilian government announced a temporary ban on foreign air travelers by March 31, 2020. Moreover, most State governors have imposed quarantines to contain the spread of the virus, but with no support from the federal government. A recent study suggests that more than 40% of social isolation in Brazil is necessary to flatten the epidemic curve of the new coronavirus and to prevent the collapse of the healthcare system (Croda et al., 2020). We defend that the only feasible way to achieve this target is to keep on with social distancing and avoid gatherings of people.

Despite the aforementioned efforts, most of them being contemporaneous among many countries, the effective containment of COVID-19 is still a delicate task because of the characteristic mild symptoms and the transmission before the full onset of the disease (Fraser et al., 2004; R. Li et al., 2020). However, the example of the sanitary action conducted in Great China was fundamental when the contagious boundaries were discrete. In Brazil’s context, there are cities such as São Paulo, Rio de Janeiro, and other capitals such as Fortaleza and Manaus that likely act as super-spreaders of SARS-CoV-2. In a network context, these cities maximize their influence on COVID-19 spread by exporting many cases to cities nearby (Madotto and Liu, 2016). This fact highlights that these centers should be of foremost concern, especially because some of them are near (e.g. Ceará and Rio de Janeiro) or have already saturated (e.g., Manaus) their health system carrying capacity, given that the number of cases is growing very fast (Crokidakis, 2020). Therefore, the multiple potential Wuhan-alike regions in Brazil require government-level actions towards the necessity of multiplicative movement restrictions and social distancing. Moreover, there are speculations that the number of documented Brazilian cases is likely more than ten times lower than the real number (Bastos and Cajueiro, 2020), considering that tests are being conducted mostly only in cases of severe acute respiratory syndrome (SARS). As an alternative to contour this situation, especially in those countries where several undocumented cases are expected (e.g., in Brazil), decision-makers could base their actions on the recent increase in number of SARS-related hospital records as a surrogate to the real amount of COVID-19 cases.

In addition to social distancing and restrictions on mobility, interventions available include rapid diagnosis and isolation of confirmed cases (Kraemer et al., 2020). However, in Brazil, the diagnosis is completely biased towards people already at advanced clinical stages, in contrast to other countries such as China, Singapore, Germany, and the United States. Clearly, the growth rates of infections by the new coronavirus differ across countries (Coelho et al., 2020; Ficetola and Rubolini, 2020). As the rate differs, the factors determining the expansion of the infection within populations inevitably result from the policies adopted by each country. Our results reveal that this variation also occurs within Brazil. We believe that the reasons for this can be addressed to the continental-wide nature of our country, or the non-uniform demographic distribution across regions (Reis-Santos et al., 2013). Thus, the responsibility for the local spread of COVID-19 is directly dependent on Mayors’ and Governors’ positions.

Several studies have shown that environmental factors such as local temperature and precipitation may affect the SARS-CoV-2 virus survival and transmission, with significant consequences for the seasonal and geographic patterns of outbreaks (Bukhari and Jameel, 2020; Ficetola and Rubolini, 2020; Ma et al., 2020). The mechanism underlying these patterns of climate determination is likely linked with the ability of the virus to survive external environmental conditions prior to reaching a host (Harmooshi et al., n.d.). A recent study showed that COVID-19 is more viable at lower temperatures (5-11°C) and was inversely related to humidity (Sajadi et al., 2020). However, our results indicate that climate variables had secondary roles in explaining the COVID-19 growth rate in Brazil. This weak relationship suggests that seasonal climatic variation plays a minor role in the spatial spread and severity of COVID-19 outbreaks, as observed in recent studies on other regions (Baker et al., 2020; Coelho et al., 2020).

For instance, Manaus, the largest metropolitan city in North Brazil, is located at the core world’s largest rainforest, the Amazon, and is characterized by extremely high rainfall and temperatures throughout all over the year. This city registered more than one thousand confirmed cases and, recently, the state health department is suffering from the collapse of the health network due to the excessive cases of COVID-19. Among these, over 85 cases have been recorded in indigenous peoples and four deaths have been confirmed, including young people. Historically, due to the absence of antibodies, indigenous people are likely more susceptible to diseases such as flu, measles, rubella, and tuberculosis, which caused dramatic epidemic cases with the arrival of Europeans on the South American continent (Montenegro and Stephens, 2006). According to the latest census, more than 300 thousand of indigenous people currently live in the Amazon region, which is home to more than 100 tribes that are still isolated. The advance of illegal mining is a major threat to these peoples. Furthermore, instead of allowing religious missionaries to get in contact with isolated Indigenous groups, all means of transportation to these areas should be restricted (Ferrante and Fearnside, 2020). Therefore, the thoughtlessness by the federal government about COVID-19 potentially leads to a dramatic scenario for indigenous people.

It is important to highlight that the low predictability of climatic variables in explaining the virus outbreaks patterns may be explained by the climatic peculiarities of Brazil during the summer that matched with the arrival and spread of the virus. Among all cities with confirmed cases, the average temperature was 24°C and the lowest temperature was 18°C, which is still far from what was indicated as ideal for the survival of the virus (Sajadi et al., 2020; Wang et al., 2020). We found a positive relationship between temperature and precipitation variables and the growth in the number of COVID-19 cases, which is in line with previous findings in Brazil (Auler et al., 2020), but contrary to a global tendency. Although the coefficients were quite weak, this suggests that high temperatures and precipitation are not limiting factors for the spread of the virus. Conversely, a scenario of extreme concern is emerging, since the Winter season approaches, so temperatures will drop dramatically across many areas, especially in the South region. Therefore, at this early outbreak, any generalization is hasty and results have to be considered with extreme caution. Most importantly, the climate must not be used as an easing argument as particularly declared by the Brazilian President.

The actual emergence of the novel infectious agent has revealed the vulnerability of societies to new health threats (Morse et al., 2012). Brazil has recently experienced other public health emergencies under polio, smallpox, cholera, H1N1 (Influenza A), avian flu, yellow fever, dengue and zika (Croda et al., 2020). Along with the current COVID-19 outbreak, we judge that all these examples constitute an important legacy on the role of scientific research on dealing with epidemics. Thus, we may now face the most important event, in the recent decades, to learn on how to respond to emergencies effectively (Croda et al., 2020).

We need to assume that this study does not intend to serve as specific guidelines for any decision-making process within any specific administrative council. Nevertheless, at a country-scale, we disapprove any government declaration or exposure that confronts the statements and protocols from the World Health Organization (WHO) or the United Nations (UN), which are international institutions that are presumed to be sovereign in their actions and recommendations since their utmost concern is the health and human wellbeing on a global scale. If eventually confronted, the arguments for doing so should be based on clear and strong scientific evidence, although it is very unlikely that these organizations have ignored such information while developing their protocols.

It is now clear that the most likely determinant of the virus spread is the vicinity of infected peoples and cities. While evacuating cities is not reasonable at this point, the easiest and the most feasible way to decelerate the COVID-19 spread is to avoid people transportation among them. This is fundamental within cities. While the virulence of the SARS-CoV-2 virus is remarkably high and we are not able to tear houses or buildings apart, the only way to allow for the healthcare systems to treat ill people effectively is performing social distancing. In the absence of any effective treatment and vaccine, we support that social distancing is still the only feasible way to avoid the collapse of our national health system. Specially in Brazil, a ‘3rd world country’, we have few hospitals and clinics with beds and respirators that are autonomous for treating SARS, which means that if we allow it to grow fast by not maintaining restrictions in movement between cities/states and social distancing within cities, we may certainly hope for the best but expect for the worst. We assume that the indicator for the efficiency of social distancing is a conundrum because the response about its effectiveness might come only weeks after the execution of any plans. In fact, people tend to underrate social isolation because the more effective it is, the less needed it seems to be. We also admit that any delayed action can be catastrophic, both for the health and economy of any country. Nevertheless, the expeditious suspension of social distancing under the coating of restoring the economic trades and jobs likely has even more dangerous side effects than COVID-19 itself.

## Data Availability

All relevant ethical guidelines have been followed; any necessary IRB and/or ethics committee approvals have been obtained and details of the IRB/oversight body are included in the manuscript.

## Acknowledgements

This study was developed as a contribution to the Brazilian Ministry of Health about the overall evaluation of collected data on COVID-19. Most of the merit of this study belongs to all those individuals involved in collecting epidemiologic data and working in favor of health, directly or indirectly, inside or outside the healthcare facilities. They deserve sincere acknowledgements for their daily work.

## Author contribution

MTB conceived the study with extensive suggestions from FMLT. MTB gathered and organized information and FMLT analyzed data. Visual results were produced by FMLT. MTB and FMLT wrote the first draft, which was thoroughly reviewed and approved by all authors.

